# Risk of Delayed Percutaneous Coronary Intervention for STEMI in the Southeast United States

**DOI:** 10.1101/2024.07.11.24310307

**Authors:** Maxwell C. Messinger, Nicklaus P. Ashburn, Joshua S. Chait, Anna C. Snavely, Siena Hapig-Ward, Jason P. Stopyra, Simon A. Mahler

**Author notes:** **Corresponding Author:** Maxwell C. Messinger Department of Family Medicine and Community Health University of Massachusetts Chan School of Medicine 119 Belmont St Worcester, MA 01605, USA.

## Abstract

**Background:** Emergent reperfusion by percutaneous coronary intervention (PCI) within 90 minutes of first medical contact (FMC) is indicated in patients with ST-segment elevation myocardial infarction (STEMI). However, long transport times in rural areas in the Southeast US make meeting this goal difficult. The objective of this study was to determine the number of Southeast US residents with prolonged transport times to the nearest 24/7 primary PCI (PPCI) center.

**Methods:** A cross-sectional study of residents in the Southeastern US was conducted based on geographical and 2022 5-Year American Community Survey data. The geographic information system (GIS) ArcGIS Pro was used to estimate Emergency Medical Services (EMS) transport times for Southeast US residents to the nearest PPCI center. All 24/7 PPCI centers in North Carolina, South Carolina, Georgia, Florida, Mississippi, Alabama, and Tennessee were included in the analysis, as well as nearby PPCI centers in surrounding states. To identify those at risk of delayed FMC-to-device time, the primary outcome was defined as a >30-minute transport time, beyond which most patients would not have PCI within 90 minutes. A secondary outcome was defined as transport >60 minutes, the point at which FMC-to-device time would be >120 minutes most of the time. These cutoffs are based on national median EMS scene times and door-to-device times.

**Results:** Within the Southeast US, we identified 62,880,528 residents and 350 PPCI centers. Nearly 11 million people living in the Southeast US reside greater than 30 minutes from a PPCI center (17.3%, 10,866,710, +/- 58,143), with 2% (1,271,522 +/- 51,858) living greater than 60 minutes from a PPCI hospital. However, most patients reside in short transport zones; 82.7% (52,013,818 +/- 98,741). Within the Southeast region, 8.4% (52/616) of counties have more than 50% of their population in a long transport zone and 42.3% (22/52) of those have more than 90% of their population in long transport areas.

**Conclusions:** Nearly 11 million people in the Southeast US do not have access to timely PCI for STEMI care. This disparity may contribute to increased morbidity and mortality.

## INTRODUCTION

Timely percutaneous coronary intervention (PCI) is the standard treatment for ST-elevation myocardial infarction (STEMI) and is associated with improved morbidity and mortality. Every 30-minute delay in reperfusion is associated with a 7.5% increase in one-year year mortality.^1^ American College of Cardiology Foundation/American Heart Association (ACCF/AHA) guidelines recommend that first medical contact (FMC)-to-device time be 90 minutes or less and, when PCI is not possible within 120 minutes of FMC, that fibrinolytic therapy be administered as early as possible.^2,3^ Traditional measures to decrease time to reperfusion include emergency medical services (EMS) recognizing STEMI rapidly, minimizing scene time, and providing early notification and 12-lead electrocardiogram (ECG) transmission to receiving facilities.^4–9^

While advances in STEMI care have led to decreased mortality rates in the US since the mid- 1980s, those in urban areas have benefitted much more than their rural counterparts.^10–13^ Rural residents have a 40% higher prevalence of cardiovascular disease (CVD) as well as higher CVD mortality – the so-called “rural mortality penalty.”^12^ The effects of the urban-rural divide are most pronounced in the American South, especially in Appalachia and the Mississippi Delta region, where the lowest life expectancies and the highest CVD mortality rates in the country are found.^10^ In response to this disparity, in 2020 the AHA announced a “Call to Action” to better understand challenges in rural cardiovascular care and to improve outcomes.^14^

Patients living in rural areas, particularly those living more than a 30-minute drive from a primary PCI (PPCI) center are less likely to receive timely PCI. To better characterize this rural cardiovascular care disparity, this study aims to quantify the risk of delayed PCI in the Southeastern US by estimating the population that would be likely to experience delayed PCI due to transport time to their nearest PPCI center. The primary objective was to quantify those residents who are unlikely to be able to receive PCI within 90 minutes of FMC. Secondary objectives were to 1) identify to what extent long transport times coincide with rural populations, 2) quantify the proportion of residents who would be unlikely to receive PCI within 120 minutes of FMC, and 3) identify the counties that have greater than half of their population at high risk of an FMC-to-device time longer than 120 minutes. These findings will help identify where the population at risk of delayed PCI resides and may help inform healthcare policymakers and prehospital care protocols.

## METHODS

### Study Design and Setting

We conducted a cross-sectional study of all residents of the Southeast US. The Southeast US is defined here as North Carolina (NC), South Carolina (SC), Tennessee (TN), Georgia (GA), Florida (FL), Mississippi (MS), and Alabama (AL). The Strengthening the Reporting of Observational Studies in Epidemiology (STROBE) guidelines were used to direct the research and manuscript development processes.^15^ The study was reviewed by the Wake Forest IRB and determined to be exempt.

### Patient Population

The study includes all residents of the Southeast US identified using the 2022 5-year American Community Survey. In addition, the 5-year American Community Survey was also used to determine resident demographics, including their age, sex, and race. The American Community Survey is a product of the US Census Bureau that estimates characteristics of the US population using a survey sent to a sample of the US population–roughly 3.5 million addresses across all 50 states, the District of Columbia, and Puerto Rico. Responses collected across years are aggregated to generate 5-year estimates that have increased reliability, especially for sparsely populated areas and small population groups.^16^

### Transport Time Calculations

Southeast US resident ground transport times to the nearest PPCI center were calculated using the Network Analyst toolkit within the geographic information service (GIS) system ArcGIS Pro.^17^ This software calculates anticipated drive times by using road network maps and proprietary travel time estimation algorithms. Specifically, the Service Area tool was used to identify travel time zones to each PPCI center identified by the study. This tool uses a list of locations and identifies the outer boundary of an area, within which the locations can be reached within a specified time. The tool was configured to estimate the drive time of emergency vehicles in rural settings to the nearest PPCI center.

To assess the accuracy of the ArcGIS Network Analyst system methodology for estimating transport time, we also generated the 30- and 60-minute travel time zones using two other mapping software packages (Here and Mapbox using the *hereR* and *mapboxapi* packages).^18–20^ We then used the outputs of each to generate independent estimates of the population at risk and characterized the variance based on the routing tool used. This analysis indicated that ArcGIS provides an estimate that is similar to other tools, with the ArcGIS-generated estimate of the population ≥30 minutes from PPCI falling between those of Here and Mapbox.

In addition, we compared the performance of our transport time calculation with both linear distance from a PPCI center and Rural-Urban Commuting Area (RUCA) code, a measure of how rural a census tract is based on population density, urbanization, and daily commuting as determined by the 2010 Decennial Census.^21,22^ Each of these variables was used in univariate logistic regression, carried out in R, to determine how reliably they could predict whether a census tract would have a short (<30 minute) or long (≥30 minute) transport time.^18,21^ Each census tract in the region was classified as a short or long transport tract based on the transport time to the tract centroid. A 75% sample of tracts in the Southeast US was used for training with the remaining 25% used for validation. Model performance was assessed by generating a confusion matrix and κ-statistic in the *caret* package.^23,24^ These Logistic regression analyses demonstrated that our GIS-based calculations of transport time were superior to linear distance or RUCA code strategies in identifying long transport zones. Use of linear distance was similar to GIS-based transport time, but only identified 90% of long transport zones compared to GIS-based transport time. Performance of logistic regression using RUCA code as a predictor was poor compared to GIS-based transport analysis, especially in predicting long transport zones.

To stratify the risk of delayed PCI, transport times were categorized as short, long, and very long (<30, 30-60, >60 minutes, respectively). Transport time categories were based on a national EMS scene time in STEMI of 15 minutes and a national average door-to-balloon time (DBT) of 45 minutes.^25,26^ Thus, our transport time thresholds were chosen to identify the population that, when transported by ambulance, would be likely to meet the ACCF/AHA goal of PCI within 90 minutes of FMC, those who will likely receive PCI within 120 minutes, and those who are unlikely to receive PCI within 120 minutes, when added to an average EMS scene time and DBT (Figure 1).^2^ For example, an EMS transport time of 30-60 minutes when added to an average scene and DBT time is likely to result in an FMC-to-device time of 90-120 minutes, indicating that these patients are unlikely to meet the 90 minute FMC-to-device time goal. A transport time >60 minutes when added to scene and DBT times is likely to result in an FMC-to-device time >120 minutes, the point at which ACCF/AHA guidelines recommend prioritizing early fibrinolytic therapy.^2^

**Figure 1:**
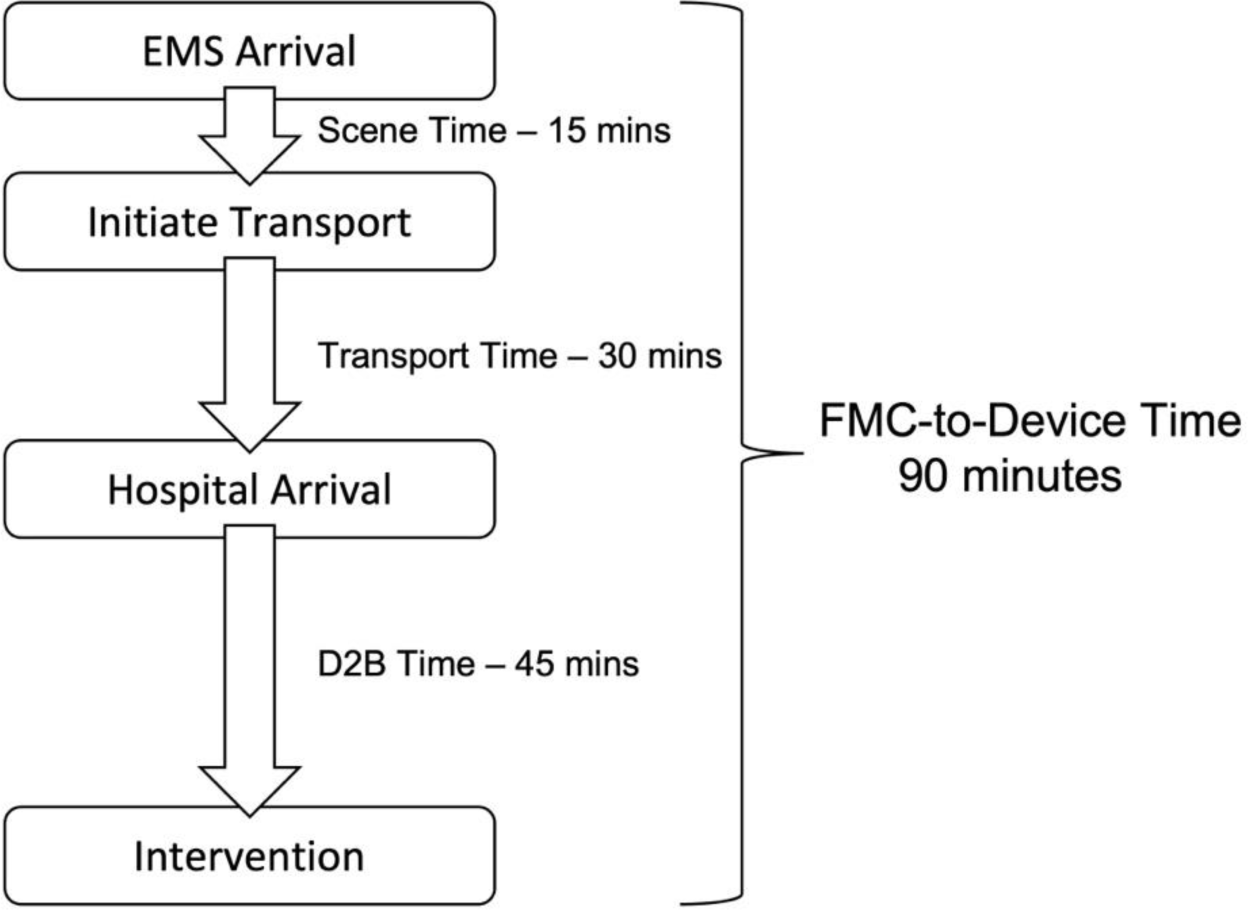
The components of first medical contact (FMC)-to-device time. The 30- and 60-minute transport time cut points were selected based upon a 15-minute scene time and a 45-minute door-to-balloon (D2B) time.

### Rurality

Rural areas were defined based on the Federal Office of Rural Health Policy (FORHP).^22^ Rural counties and census tracts were used to determine the percentage of long-transport areas contained in rural areas as well as the percentage of rural areas that do not have a high risk of long transport. The degree of rurality for short and long-transport areas was also calculated using RUCA codes.^21^

### Primary PCI Center Identification

The CathPCI Registry, published by the American College of Cardiology (ACC), was used to identify facilities performing PCI in the Southeast US.^27,28^ This database relies on voluntary participation by cardiac catheterization labs (CCLs) but includes more than 90% of CCLs in the US.^29^ To identify PCI centers that were available 24 hours a day and 7 days a week, the study team called each facility and spoke with the CCL staff using a standardized call script (Supplemental Appendix 1). Calls were made between June 2021 and April 2022. If CCL staff were unavailable, team members spoke with the emergency department charge nurse or attending emergency physician. Every CathPCI-registered CCL in the Southeast US was called. Additionally, study team members called select nearby CCLs in surrounding states (Virginia, Kentucky, Missouri, Arkansas, and Louisiana) to determine if they were PPCI centers. Lastly, the number of PCI centers within FORHP-defined rural areas was calculated.

### Delayed PCI Risk Analysis

The total number of residents within each transport time category, along with the 95% margin of error, was estimated using the R Statistical Computing Environment, along with the *tidyverse* package and data from the 2022 5-year American Community Survey.^18,30–32^ Population data at the census tract and block level were used to identify the population within each transport zone. To handle tracts that lie across multiple transport categories, a population-weighted interpolation method from the *tidycensus* package was used to assign a proportion of the tract’s population to each zone.^33^ This approach uses the estimated population at a finer scale, in this case the block level, as a weight to assign a population to a subset of a larger geometry, in this case the tract.^34^ The total population within each transport zone was then calculated along with the proportion of patients living <30 minutes from PPCI, 30-60 minutes from PPCI, and >60 minutes from PPCI. Patient demographics including age, sex, and race of the population residing within the Southeast US were summarized using descriptive statistics, and 95% margin of error was estimated using *tidycensus*.^33^ Differences in these demographics between transport zones were evaluated using two-tailed z-tests as described in the American Community Survey guidance document.^35^

## RESULTS

Within the Southeast US, we identified a total population of 62,880,528 +/- 102,975 people, of which 51.0% (32,062,915 +/- 62,809) were female, 63.6% (39,967,520 +/- 82,264) were white, and 43.7% (27,459,924 +/- 52,606) were age 45 or older. We identified 350 PPCI centers within the Southeast US and 23 PPCI centers in adjacent states (Figure 2). Florida is home to the most PPCI centers in the region with 150 (42.9%) while Mississippi is home to the least with 22 (6.3%). While most PPCI centers were in urban areas, 9.4% (33/350) were in FORHP-defined rural areas.

**Figure 2:**
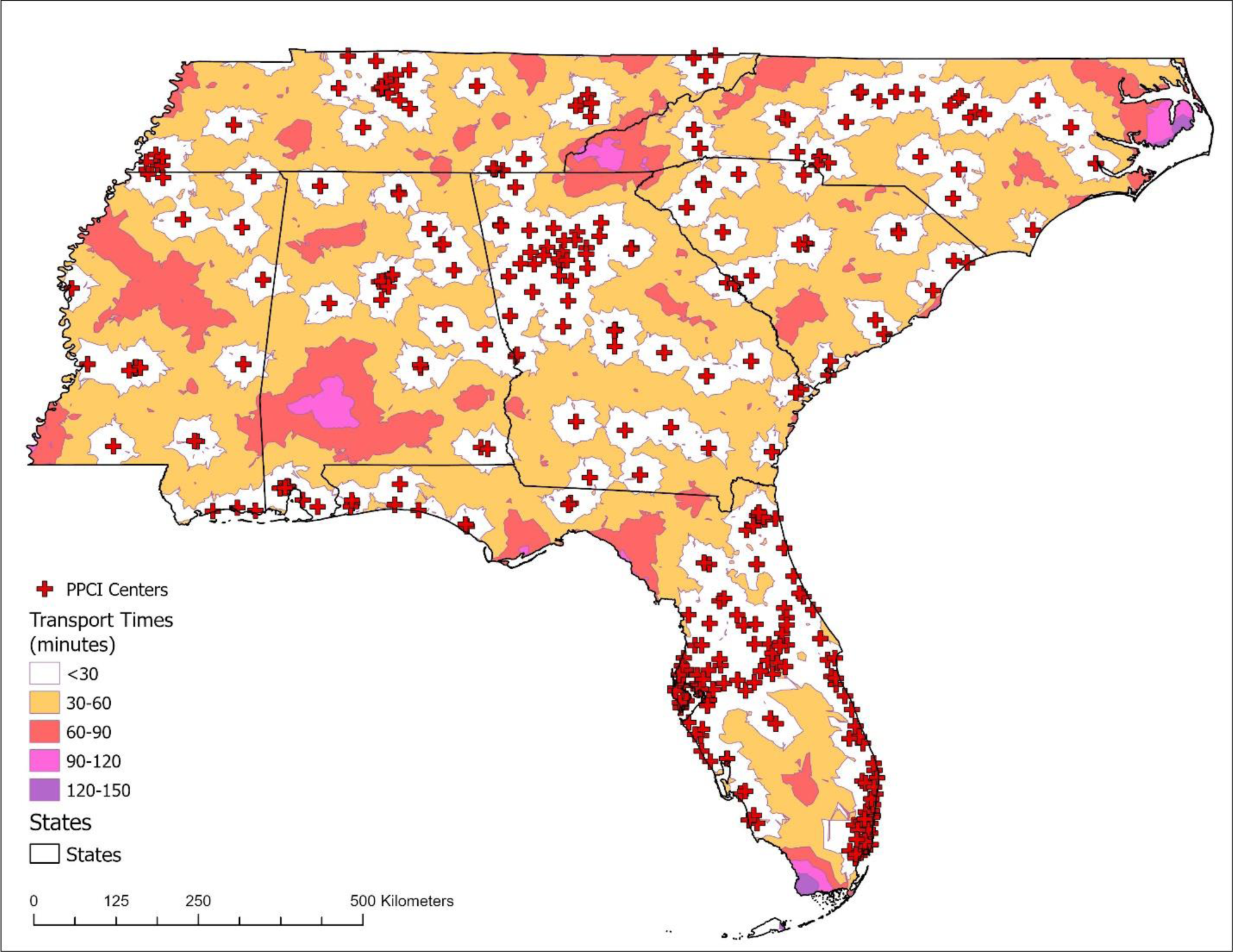
Primary percutaneous coronary intervention (PCI) centers and transport time zones in the Southeast US. Five different transport time regions are shown, demonstrating the range of transport times that exist within the region.

Within the Southeast US population, 17.3% (10,866,710 +/- 58,143) had an estimated transport time to PPCI ≥30 minutes, corresponding to a high likelihood of an FMC-device time >90 minutes. Among these patients, most have a transport time within 30-60 minutes, but 11.7% (1,271,522 +/- 51,858) reside >60 minutes from the nearest PPCI center, which makes them unlikely to have an FMC-to-device within 120 minutes. Therefore, 2.0% (1,271,522 +/- 51,858) of the total population in the Southeast US is likely to have an FMC-to-device time of greater than 120 minutes. Within the Southeast US, 82.7% (52,013,818 +/- 98,741) of the population live less than 30 minutes from PPCI and are at low risk of delayed PCI due to geography (Table 1).

**Table 1:** Demographic and socioeconomic characteristics of residents of the Southeast US. All data collected from the 2020 5-Year American Community Survey. Parentheses contain 95% margin of error estimate.

|  | **Southeast US** | **≤30 Minute Transport** | **>30 Minute Transport** |
| --- | --- | --- | --- |
| Total Population | 62,880,528 (102,975) | 52,013,818 (98,741) | 10,866,710 (58,143) |
| Race |  |  |  |
| White | 39,967,520 (82,264) | 32,246,852 (78,267) | 7,720,668 (46,755) |
| Non-White | 27,754,410 (99,221) | 24,093,141 (96,705) | 3,661,269 (96,703) |
| Sex |  |  |  |
| Female | 32,062,915 (62,809) | 26,625,963 (60,239) | 5,436,952 (32,892) |
| Age |  |  |  |
| <25 | 19,261,074 (51,543) | 15,983,749 (49,176) | 3,277,325 (48,900) |
| 25-44 | 16,159,530 (45,102) | 13,592,642 (43,180) | 2,566,888 (42,883) |
| 45-64 | 16,198,293 (41,416) | 13,305,734 (39,460) | 2,892,559 (39,071) |
| ≥65 | 11,261,631 (32,437) | 9,131,693 (30,764) | 2,129,938 (30,319) |

Among patients at risk of delayed PCI due to long or very long transport times, 71.0% are white (7,720,668 +/- 46,755) compared to 62.0% (32,246,852 +/- 78,267) in short transport zones (p<0.0001). We also found that the population in long transport areas is older, with 46.2% (5,022,498 +/- 49,455) of the population ≥45 years old in long transport areas compared to 43.1% (22,437,426 +/- 50,035) in short transport areas (p<0.0001).

### Areas at Risk

Long transport zones are distributed across the entire Southeast US and are present to varying degrees in every state (Figure 2). Areas with the longest transport times are in the mountains of NC, the NC coast, the FL panhandle, the FL keys, and portions of AL and MS (Figure 2).

Transport times in some of these areas exceed two hours.

Most counties in the Southeast US (56%, 346/616) have >50% of their population residing greater than 30 minutes from PPCI, indicating that most counties have a large amount of their population at risk of delayed PCI (Supplemental Figure 1). Further, there is a subset of counties (8.4%, 52/616) that have >50% of their population residing > 60 minutes from PPCI. Of those, 42.3% (22/52) have more than 90% of their population living over 60 minutes from PPCI (Figure 3).

**Figure 3:**
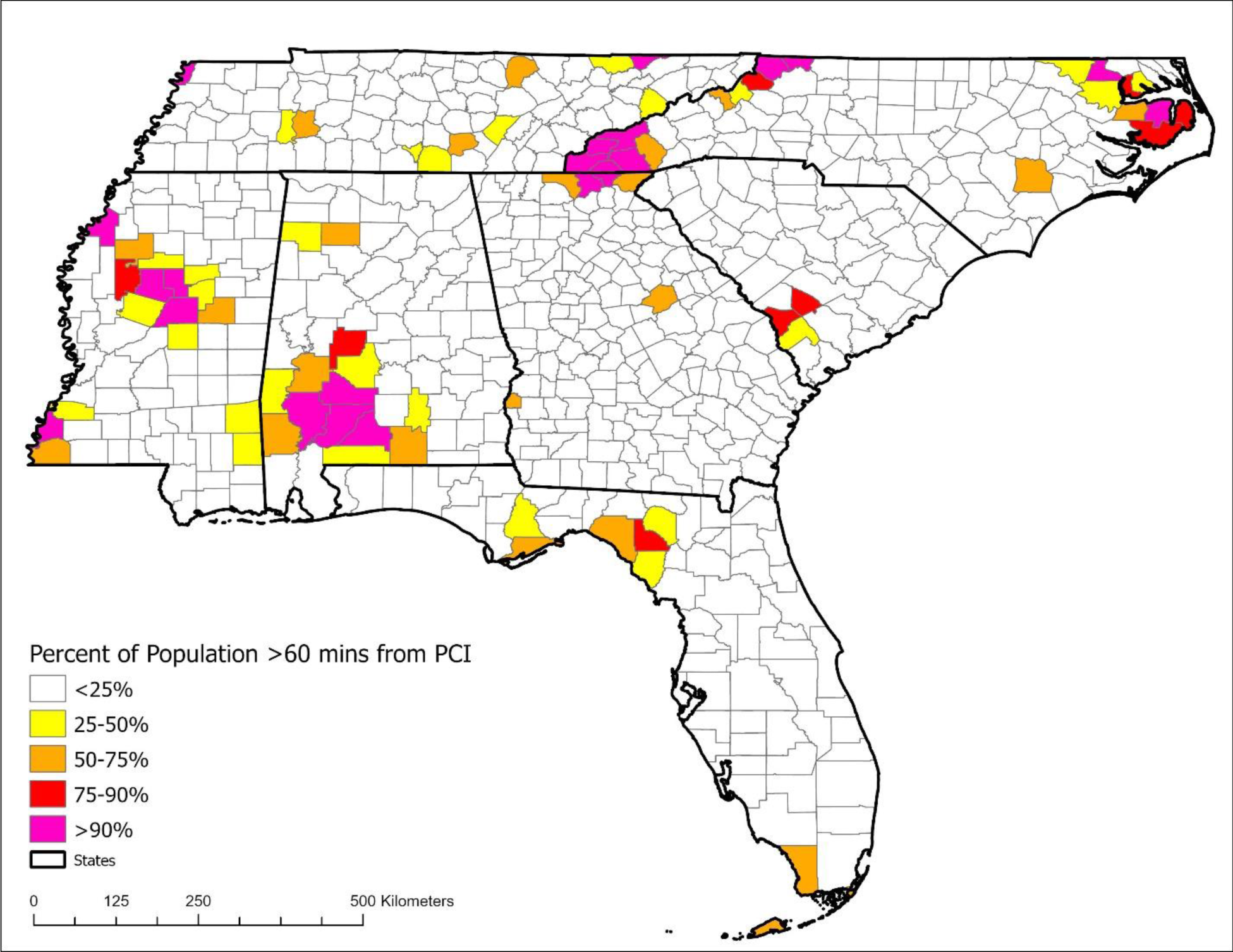
Counties of the Southeastern US by the proportion of the population that resides greater than 60 minutes from a primary percutaneous coronary intervention (PCI) center. Those living greater than 60-minute from primary PCI are unlikely to receive intervention within 120 minutes of first medical contact (FMC).

### Rural Areas and Transport Times

Most areas that are ≥30 minutes from PPCI in the Southeast US are contained within a rural area (85.6%). While long transport zones are mostly contained in rural areas, 37.7% of rural areas have transport times <30 minutes from PPCI (Supplemental Figure 2). Long transport zones are also likely to be more rural than short transport zones as determined by RUCA code. Short transport zones have a mean RUCA of 1.79 (SD 4.24) while long transport zones have a mean RUCA of 4.40 (SD 4.23). Those areas with a RUCA of 4 or higher are classified as rural by the FORHP, indicating that the average long transport area is rural according to the FORHP definition.

## DISCUSSION

This is the first study to investigate the population at risk of delayed PCI in an entire geographical region. We found that nearly 11 million people in the Southeast US reside in long transport zones and are at high risk for delayed PCI. Of those, over 1.2 million people reside more than 60 minutes from PPCI, meaning that it is unlikely they would reach PCI within 120 minutes of FMC when transported by ground. In these patients, alternative transports methods, such as helicopter EMS, or alternative interventions, such as prehospital fibrinolysis, should be considered. Additionally, we found that people in western NC, the NC coast, the FL Keys and Everglades, and some areas of AL would experience ground transport times greater than two hours, making delayed PCI almost certain. These findings suggest that novel care strategies and approaches are needed to ensure equitable cardiovascular care for this at-risk rural population.

The present study is the first to use a GIS approach at a regional scale, as all previous studies have limited their scope to the state level. Our results are generally consistent with those studies investigating geographic limitations on access to PCI at the state level. Studies that examined geographic limitations on access to PCI in Alabama, Ohio, and Maine, found that there is an association between rural residency and delayed access to cardiac interventional services.^36–38^ However, most of these studies were limited by the proxies for travel time that were used, such as linear distance to a PPCI center. Our data suggest that linear distance from tract centroids to PPCI predicts transport time well but, since this method collapses the geography of tracts to their centroids, it may not provide an adequate characterization of census tracts that extend across multiple transport time zones. This limitation may become more important in larger rural tracts, the largest of which in the Southeast US is 2,410 square km, or roughly 75% the size of Rhode Island. Graves used a similar GIS approach to the one used in the present study, but this analysis was limited to a single state.^38^

Our results add to a growing body of literature establishing an important rural care disparity for STEMI care. To address this disparity, policymakers need data to determine where the greatest needs exist for increased cardiovascular services. Most PPCI centers are mostly found in areas of high population density where they can expect sufficient volumes to support their operations. Studies have shown that while many PCI centers do not meet case volume targets, which may have a small adverse effect on outcomes, opening PCI centers does lead to an increase in rates of same-day PCI for acute coronary syndrome.^39,40^ Knowing this, policymakers should determine whether PCI facilities can be located in areas of geographic need.

In areas where adding PCI facilities is not feasible, there must be renewed efforts to ensure that patients can be transported rapidly across large distances and treated quickly upon arrival at a hospital. Prehospital ECG transmission and activation of the CCL before patient arrival, an approach adopted by many hospitals already, has been shown to reduce FMC-to-device time and decrease mortality.^41,42^ Additionally, transport by helicopter emergency medical services (HEMS) could provide a reduction in transport time. In certain scenarios, HEMS has been shown to significantly reduce transport times, though this may not always lead to an FMC-to-device time <90 minutes.^43–45^ The current study could aid EMS agencies in protocolizing HEMS as a transport mode for STEMI. Finally, early prehospital administration of fibrinolytic therapy or diversion to the closest fibrinolytic-capable emergency department may provide a solution in patients unlikely to receive PCI within 120 minutes. Prehospital fibrinolysis has been adopted in many rural areas of the world and is seeing increased adoption in the US.^46–48^ Further adoption of this approach may be facilitated through emerging technologies such as telemedicine.

### Limitations

The current study did not consider the use of air transport. This would reduce transport times, but HEMS availability is affected by factors such as inclement weather and a limited number of aircraft. As a result, our analysis provides useful information about what transport times can be achieved in any conditions. Estimates of ground transport time are also imperfect, although previous comparisons of ArcGIS analyses and documented real-world transport times indicate that the software tends to underestimate transport time by approximately 12 minutes when transport time exceeds 20 minutes.^49^ Therefore, our results are likely a conservative estimate of the population at risk from long ground transport. Additionally, US Census data is subject to sampling bias and an incomplete survey response rate.^50^ The area-weighted approach used to assign population estimates to block groups including multiple transport zones is also imperfect, as it assumes that the population is evenly distributed across each block group. We do not, however, expect that these assumptions would introduce a systematic bias in the results. For this study, we relied on the CathPCI registry to provide a list of facilities that perform PCI from which PPCI centers were identified. While the CathPCI registry does not include all CCLs in the country, it does include greater than 90% and we expect that participation among PPCI centers is likely to be higher than among all CCLs.^29^ Regardless, some PPCI centers may not be included in the current study. We also relied on calls directly to facilities to identify PPCI centers and the knowledge of hospital staff. It is possible these methods underestimated or overestimated the number of PPCI centers.

## CONCLUSIONS

Nearly 11 million people in the Southeast US are at risk of delayed PCI for STEMI. There are 1.2 million residents of the Southeast US who live greater than 1 hour from PPCI. This cardiovascular care disparity places these patients at risk of increased morbidity and mortality from STEMI and intersects with other rural health disparities to contribute to worse CVD outcomes. Policymakers, health system leaders, and prehospital medical directors should use this information to help develop equitable care strategies regarding primary PCI center development and location, further optimization of door-to-device time for these patients, as well as novel prehospital care approaches, such as EMS-based fibrinolysis protocols and protocolized air transport for STEMI.

## Data Availability

Data and R script used for analysis available from the corresponding author upon request.

## Acknowledgments

We thank those who made this project possible, especially Alex Ambrosini, Allison Perko, Ching-Hsuan Lin, Ian Kinney, Jake Nearine, Justin Holbrook, Mark Perez, Michael Chado, and Robert Ledbetter, who volunteered their time to collect the data on which the current study relies.

## Previous Presentations

Preliminary data were presented at Wake Forest Emergency Medicine E. Byrum Academic Forum in Winston-Salem, NC on April 20, 2022 and the Society for Academic Emergency Medicine Annual Meeting in Austin, TX on May 18, 2023.

